# Ongoing long-lasting insecticide-treated net distribution efforts are insufficient to maintain high rates of use among children in rural Uganda

**DOI:** 10.1101/2021.02.26.21252527

**Authors:** Claire M. Cote, Varun Goel, Rabbison Muhindo, Emmanuel Baguma, Moses Ntaro, Bonnie E. Shook-Sa, Raquel Reyes, Sarah G. Staedke, Edgar M. Mulogo, Ross M. Boyce

**Author notes:** Corresponding Author: Ross M. Boyce, Division of Infectious Diseases, University of North Carolina at Chapel Hill, 130 Mason Farm Road, CB 7030, Chapel Hill, NC 27599.

## Abstract

**Background:** Long-lasting insecticide-treated nets (LLINs) remain a cornerstone of malaria control, but optimal distribution strategies to sustain universal coverage are not well-defined

**Methods:** We conducted a cross-sectional survey of 2,190 households in the highlands of western Uganda to examine LLIN source and use among children age with elevation and distance to clinic being the primary variables of interest.

**Results:** We found that only 64.7% (95% CI 64.0 – 65.5%) of children were reported to have slept under a LLIN the previous night. Compared to those living <1 km from a health center, households at ≥ 2 km were less likely to report the child sleeping under a LLIN (RR 0.86, 95% CI: 0.83 – 0.89, *p*<.001). Households located farther from a health center received a higher proportion of nets from government distributions compared to households living closer to health centers.

**Conclusions:** Continuous, clinic-based distribution efforts were insufficient to sustain high rates of LLIN use among children between mass distribution campaigns. More frequent campaigns and complementary approaches are required to achieve and maintain universal LLIN coverage in rural areas.

## BACKGROUND

Malaria remains an important cause of global morbidity and mortality despite substantial gains against the disease over the past two decades [1]. Much of the progress against malaria can be attributed to the development and widespread implementation of long-lasting insecticide-treated nets (LLINs) [2]. When widely distributed in the community and employed in the household, LLINs provide both a physical barrier against the bite of female *Anopheles* mosquitoes as well as a killing effect (i.e., vector control) resulting from contact between the mosquito and the impregnated pyrethroid insecticide [3]. Yet the emergence of resistance to pyrethroid insecticides, including permethrin and deltamethrin, threatens many of these gains [4]. Recent reports suggest that global progress against malaria has stalled and may even be slipping backwards among high-burden countries in sub-Saharan Africa (SSA) [1]. Nets employing novel insecticides or combinations of insecticides have shown to be effective in settings with established insecticide resistance, but these nets are not yet widely deployed [5, 6]. Therefore, continued focus on the development of effective implementation strategies to achieve universal coverage, which the World Health Organization (WHO) defines as one LLIN for every two persons at risk of malaria, remains a critical undertaking [7].

Among malaria-endemic countries in SSA, Uganda has been a leader in the effort to achieve universal coverage [8]. Uganda conducted its first mass LLIN distribution campaign in 2013, with over 20 million nets distributed [9]. This effort was followed by similar campaigns every three years, including in 2017-18 and most recently in 2020-21 in accordance with WHO guidelines [7]. Households reporting at least one LLIN increased from 16% in the 2006 Demographic and Health Survey (DHS) to more than 80% in the 2018 Malaria Indicator Survey, while over the same period the proportion of households with at least one LLIN for every two people increased from 5% to 54% [10]. Furthermore, in the years immediately following the initial distribution campaign, substantial reductions in malaria parasite prevalence and disease burden were observed [11]. Towards the end of each three-year cycle, however, attrition due to physical damage and even loss of nets can leave households well below universal coverage targets with a resulting increase in malaria transmission intensity [12, 13].

To maintain coverage between mass distribution campaigns, the WHO recommends continuous LLIN distribution through antenatal care clinics and the expanded program on immunization. These channels, which leverage public health services utilized by at-risk populations (e.g., pregnant women and young children), aim to fill coverage gaps that emerge due to population growth in the interval period between mass distribution campaigns. However, strategies to replace nets that experience premature attrition are not as well-defined. This may be partly attributable to the high cost of monitoring LLIN durability and performing gap analysis [14, 15]. At present, the WHO does not recommend replacement or “top-up” campaigns because “accurate quantification for such campaigns is generally not feasible and the cost of accounting for existing nets outweighs the benefits [7].”

A much smaller proportion of the existing literature has examined the effectiveness of LLIN distribution outside of mass distribution campaigns, [16-21] particularly in regard to geographic factors that may impact the coverage. While analysis of routine DHS data from 25 countries found that facility-based distribution improves LLIN ownership rates and reported use [22] a study in rural Kenya found that increased distance from health facilities was associated with decreased bed net ownership [23] and another a study in Malawi found that households further from health facilities were less likely to own a net and have their child sleep under it [24] Therefore, as part of an ongoing, cross-sectional study of malaria transmission in the western Ugandan highlands, we sought to examine how geographic factors, including elevation and distance to clinic, might influence malaria risk and LLIN use in order to inform future distribution strategies.

### Study Site

The Bugoye sub-county, located in the Kasese District of Western Uganda is comprised of 35 villages, spanning a rural, highland area of approximately 55 km^2^. The population of the sub-county is 50,249, approximately one-quarter of whom are children under five years of age [15]. The geography of the sub-county is highly varied and characterized by deep river valleys and steep hillsides with elevations up to 2,500 meters (**Figure 1**).

**Figure 1:**
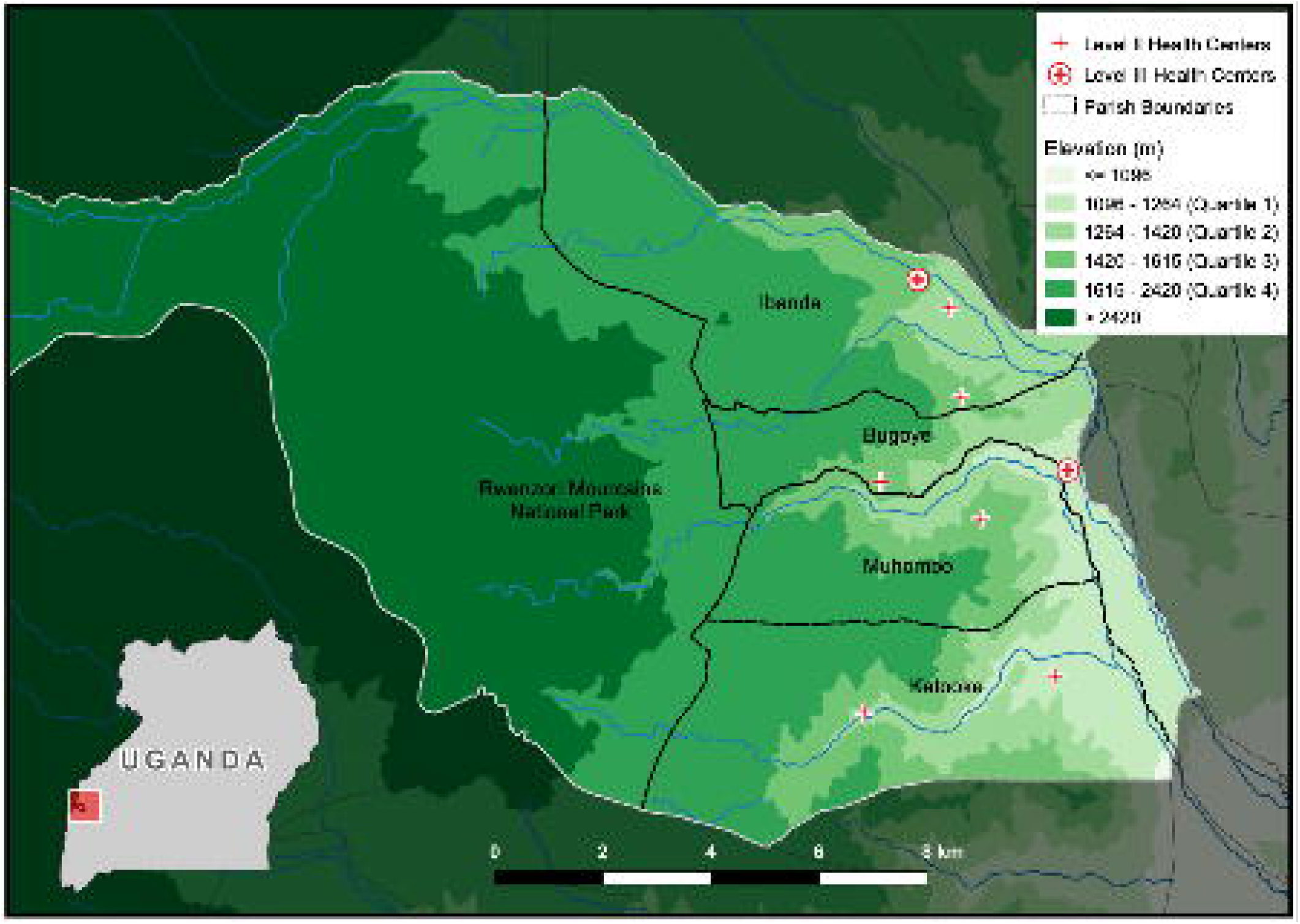
Elevation map of Bugoye sub-county displaying parish boundaries and location of Level II and Level III health centers.

The sub-county’s primary public health facility is the Bugoye Level III Health Center (BHC). BHC is comprised of a 25-bed inpatient ward, where patients can receive intravenous medications, a busy outpatient clinic that evaluates 60-80 patients per day, a maternity ward, and a small laboratory capable of performing point-of-care tests for diseases such as malaria and HIV. There are also level II health centers in each of the six parishes that offer basic outpatient services including routine vaccination, and one private-not-for-profit level III health center operated by the Rwenzori Mountaineering Services.

The climate in Bugoye permits year-round malaria transmission marked by semi-annual transmission peaks typically following the end of the rainy seasons in May and December [25]. The most recent malaria indicator surveys undertaken in the Mid-Western region (2014-15) and Tooro sub-national region (2018-19) which include Bugoye, reported *P. falciparum* parasitemia rates (PfPR) of 17.4% and 7.3%, respectively [10, 26]. The most recent mass government distribution of LLINs took place in 2017 and is supplemented by ongoing distributions through antenatal and immunization clinics.

### Household Survey

Household surveys were conducted in each of the 35 villages of Bugoye sub-county. Prior to each survey, community health workers (CHWs) disseminated information about the aims and methods of the study to the residents of their respective coverage areas in an attempt to maximize participation. During the survey, CHWs guided study staff to the nearest household with an eligible child (age 2 – 10 years) residing in the home. An adult caregiver provided written consent to participate in the study. Children ≥8 years of age were asked to provide written assent to participate. If multiple eligible children were present in the household, a random number generator was used to create an integer sequence with values between 2 and 10. The first child to have an age matching a number in the sequence was selected for testing. If there was no adult present at the time of the visit, the survey team recorded the GPS location and moved to the next household.

After consent was provided, the study team administered a brief questionnaire that elicited responses about care-seeking behaviors, bed net ownership and use, and recent health (available in Supplementary Material). Axillary temperature was measured in all children and 50 μl of capillary blood drawn for a malaria rapid diagnostic test (RDT) (SD Bioline Malaria Ag P.f., Abbott Laboratories, Chicago, IL, USA). The RDT is a qualitative test for the detection of histidine-rich protein II (HRP-II) antigen of *Plasmodium falciparum* in human whole blood [27]. RDT results were recorded as either positive or negative, with faint lines being considered positive. Results were provided to the consenting caregiver and recorded in the questionnaire.

All children with a history of fever in the prior 48 hours or documented fever (axillary temperature ≥37.5° C) at the time of initial evaluation and a positive RDT test result received weight-based treatment with artemether-lumefantrine [28]. Children with a positive RDT test result, but no reported history of fever or documented fever on initial evaluation were not treated as this may represent HRP-II antigen persistence following recent treatment or asymptomatic parasitemia, which is consistent with current national guidelines [29]. Children with fever and a negative RDT test result were referred to the nearest public health facility for further evaluation.

### Data Management & Analysis

The sample size was estimated to achieve a coefficient of variation of approximately 20% for village-level malaria prevalence estimates. Based on these calculations, we planned to survey 60 eligible households in each village - further stratified into twelve households per CHW in order to achieve spatial distribution within each village. CHWs and study staff first visited the nearest house to the CHW’s home, then moved in a clockwise direction, visiting every other household until the required number of households had been surveyed. All information was recorded in and uploaded to a secure electronic database (i.e. REDCap) using a portable tablet device [30]. Data were analyzed using Stata version 16 (College Station, Texas). After the survey was complete, data was cleaned by manual review. Minor typographical errors were corrected for temperature, latitude, and longitude. Entries without evaluable latitude and longitude were excluded from further analysis.

The following outcome measures were assessed: (i) parasite prevalence or PfPR defined as the proportion of children with a positive malaria RDT result among all tests performed (ii) LLIN use among children, measured by asking the caregiver if the participating child slept under a net the previous night, and (iii) the source of the LLIN. Weighted estimates of parasite prevalence and LLIN use were generated using the svyset command in Stata, which accounted for the estimated probability of selection for each household, sample stratification, and the finite population correction (FPC) factors [31]. Village population estimates were obtained from the most recent CHW census and were used to determine sampling weights and FPC factors. Unless stated otherwise, all estimates are weighted to the sub-county population. Weighted categorical outcomes were analyzed using Pearson’s Chi-squared test and binary outcomes were modeled using log-binomial regression to estimate crude and adjusted risk ratios (RR).

Elevation data for each household location was derived using the Google Elevation Application Programming Interface. Elevation quartiles were generated in Stata using the xtile command. Euclidean distances were calculated for both distance to nearest health center (level II or III) and distance to nearest level III health center. Distances were categorized by <1 km, 1-2 km, and >2 km to nearest health center level II or III. The association between bed net use and distance to health centers was estimated from a design-consistent log binomial regression model.

### Ethical Approvals

Ethical approval of the study was provided by the institutional review boards of the University of North Carolina at Chapel Hill (19-1094), the Mbarara University of Science and Technology (06/03-19), and the Uganda National Council for Science and Technology (HS 2628).

## RESULTS

From January 8 to March 11, 2020, field staff surveyed a total of 2,190 households, representing 31.8% of all households in the sub-county. After removal of erroneous values, 99.2% (2,173 of 2,190) of entries had evaluable GPS coordinates, while malaria rapid diagnostic test results were available for 99.9% (2,170 of 2,173) of entries. Overall, 6.8% (148 of 2,170) of children age 2 – 10 years of age had a positive RDT result, yielding a weighted estimate of 5.8% (95% confidence interval [CI] 5.4 – 6.2%). Yet, we observed substantial variability in the positivity rates among villages, ranging from 0% (0 of 360) in six villages to a high of 31.7% (19 of 60) in Kansanzi village. A summary of household characteristics and malaria positivity prevalence (e.g., PfPR) stratified by elevation quartile is shown in **Table 1**. High-elevation villages had a lower PfPR than lower-elevation villages, and a smaller proportion of children with a self-reported fever had a positive RDT at the time of the survey.

**Table 1:**
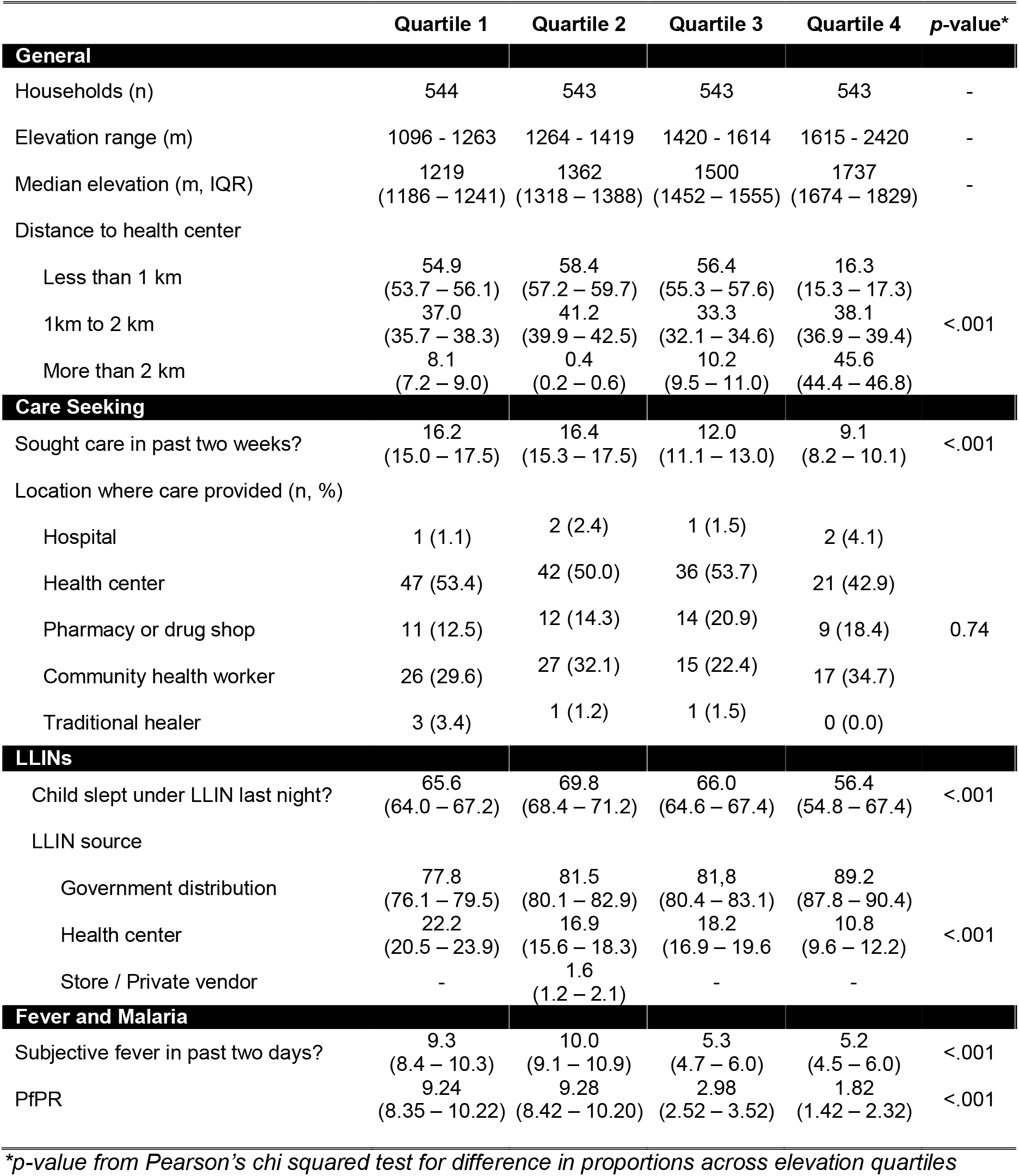
Summary of household characteristics stratified by elevation quartile. Unless otherwise indicated, data presented represents weighted proportion of households with corresponding 95% confidence intervals.

|  | Quartile 1 | Quartile 2 | Quartile 3 | Quartile 4 | p-value* |
| --- | --- | --- | --- | --- | --- |
| General |  |  |  |  |  |
| Households (n) | 544 | 543 | 543 | 543 | - |
| Elevation range (m) | 1096 - 1263 | 1264 - 1419 | 1420 - 1614 | 1615 - 2420 | - |
| Median elevation (m, IQR) | 1219<br>(1186 – 1241) | 1362<br>(1318 – 1388) | 1500<br>(1452 – 1555) | 1737<br>(1674 – 1829) | - |
| Distance to health center |  |  |  |  |  |
| Less than 1 km | 54.9<br>(53.7 – 56.1) | 58.4<br>(57.2 – 59.7) | 56.4<br>(55.3 – 57.6) | 16.3<br>(15.3 – 17.3) | <.001 |
| 1km to 2 km | 37.0<br>(35.7 – 38.3) | 41.2<br>(39.9 – 42.5) | 33.3<br>(32.1 – 34.6) | 38.1<br>(36.9 – 39.4) |  |
| More than 2 km | 8.1<br>(7.2 – 9.0) | 0.4<br>(0.2 – 0.6) | 10.2<br>(9.5 – 11.0) | 45.6<br>(44.4 – 46.8) |  |
| Care Seeking |  |  |  |  |  |
| Sought care in past two weeks? | 16.2<br>(15.0 – 17.5) | 16.4<br>(15.3 – 17.5) | 12.0<br>(11.1 – 13.0) | 9.1<br>(8.2 – 10.1) | <.001 |
| Location where care provided (n, %) |  |  |  |  |  |
| Hospital | 1 (1.1) | 2 (2.4) | 1 (1.5) | 2 (4.1) | 0.74 |
| Health center | 47 (53.4) | 42 (50.0) | 36 (53.7) | 21 (42.9) |  |
| Pharmacy or drug shop | 11 (12.5) | 12 (14.3) | 14 (20.9) | 9 (18.4) |  |
| Community health worker | 26 (29.6) | 27 (32.1) | 15 (22.4) | 17 (34.7) |  |
| Traditional healer | 3 (3.4) | 1 (1.2) | 1 (1.5) | 0 (0.0) |  |
| LLINs |  |  |  |  |  |
| Child slept under LLIN last night? | 65.6<br>(64.0 – 67.2) | 69.8<br>(68.4 – 71.2) | 66.0<br>(64.6 – 67.4) | 56.4<br>(54.8 – 67.4) | <.001 |
| LLIN source |  |  |  |  |  |
| Government distribution | 77.8<br>(76.1 – 79.5) | 81.5<br>(80.1 – 82.9) | 81.8<br>(80.4 – 83.1) | 89.2<br>(87.8 – 90.4) | <.001 |
| Health center | 22.2<br>(20.5 – 23.9) | 16.9<br>(15.6 – 18.3) | 18.2<br>(16.9 – 19.6) | 10.8<br>(9.6 – 12.2) |  |
| Store / Private vendor | - | 1.6<br>(1.2 – 2.1) | - | - |  |
| Fever and Malaria |  |  |  |  |  |
| Subjective fever in past two days? | 9.3<br>(8.4 – 10.3) | 10.0<br>(9.1 – 10.9) | 5.3<br>(4.7 – 6.0) | 5.2<br>(4.5 – 6.0) | <.001 |
| PfPR | 9.24<br>(8.35 – 10.22) | 9.28<br>(8.42 – 10.20) | 2.98<br>(2.52 – 3.52) | 1.82<br>(1.42 – 2.32) | <.001 |

\*p-value from Pearson's chi squared test for difference in proportions across elevation quartiles

Of those surveyed, 64.7% (95% CI 64.0 – 65.5%) of caregivers reported that the participating child slept under a bed net the previous night. The vast majority of respondents reported obtaining the net from either a government distribution campaign (n=1,119, 82.1%) or a health facility (n=265, 17.2%). Only four households reported purchasing a net from a vendor. The proportion of children sleeping under a net was similar in the sites of lowest elevation (Quartile 1 and Quartile 2, **Table 1**), but was lower in households at higher elevation when compared to the lowest quartiles.

Among households reporting LLIN use, an estimated 5.4% (95% CI 5.0 – 5.8%) had a positive RDT result, whereas 6.6% (95% CI 6.0 – 7.3) of children who were not reported to have slept under a net had a positive RDT result *(p*=.002). In the univariate analysis, children who reported using LLINs were less likely to have a positive RDT result compared to children who did not use nets (RR 0.83, 95% CI 0.72-0.93). At lower elevation sites, the risk of a positive RDT result was greater in children who did not use bed nets compared to those who did. However, at the highest elevation sites, where malaria transmission is presumably lowest, no difference in malaria risk was observed for children who used nets versus those who did not (**Table 2**).

**Table 2:** Results from univariate (left columns) and multivariate (right columns) log binomial regression modeling of a positive malaria RDT result

| Variable | RR | 95% CI | p-Value | aRR | 95% CI | p-Value |
| --- | --- | --- | --- | --- | --- | --- |
| **Bed Net** | 0.82 | 0.72 – 0.93 | 0.002 | 0.75 | 0.66 – 0.85 | <.001 |
| **Elevation** |  |  |  |  |  |  |
| <u>Quartile 1</u> |  | REF |  | - | - | - |
| No Net | - | - | - |  | REF |  |
| Yes Net | - | - | - | 0.65 | 0.53 – 0.81 | <.001 |
| <u>Quartile 2</u> | 1.00 | 0.87 – 1.16 | 0.96 | - | - | - |
| No Net | - | - | - | 0.87 | 0.70 – 1.09 | 0.24 |
| Yes Net | - | - | - | 0.74 | 0.61 – 0.90 | 0.002 |
| <u>Quartile 3</u> | 0.32 | 0.27 – 0.39 | <0.001 | - | - | - |
| No Net | - | - | - | 0.33 | 0.25 – 0.44 | <.001 |
| Yes Net | - | - | - | 0.21 | 0.16 – 0.28 | <.001 |
| <u>Quartile 4</u> | 0.20 | 0.15 – 0.26 | <0.001 | - | - | - |
| No Net | - | - | - | 0.15 | 0.10 – 0.23 | <.001 |
| Yes Net | - | - | - | 0.15 | 0.11 – 0.22 | <.001 |

Note: Univariate model regresses RDT result on elevation quartile, and multivariate model regresses RDT result on elevation quartile, bed net use, and their interaction. Abbreviations: CI = confidence interval; RR = risk ratio; aRR = adjusted risk ratio

To further explore the relationship between LLIN use and geographic factors, we examined rates of reported bed net use stratified by distance to the nearest health facility. In the first analysis, we estimated the shortest Euclidean (i.e., straight-line) distance to either a level II or level III facility, where bed nets are routinely provided to pregnant women seeking antenatal care and children receiving immunizations. Distance from either a level II or III health center ranged from 0.01 km (11 m) to 6.55 km with a median of 1.12 km and interquartile range 0.70 – 1.69 km. However, approximately 1 in 7 (15.2%, 95% CI 14.8 – 15.6) households was located more than 2 km from the nearest health facility. Households at lower elevations were more likely to live closer to healthcare facilities (**Table 1**). For example, at the lowest three elevation quartiles, approximately half of respondents live less than 1 km from a level II or III health center, whereas at the highest elevation quartile approximate half live ≥ 2 km away from a health center.

As shown in **Figure 2**, reported bed net use declined among households living ≥ 2 km from the nearest level II or level III facility. Compared to those living <1 km from a health center, households at more than 2 km were less likely to report the child sleeping under a LLIN (RR 0.86, 95% CI: 0.83 – 0.89, *p*<.001) (**Table 3**). We repeated the analysis using only the distance to level III facilities, which house the only labor and delivery wards and inpatient units in the sub-county. Again, we observed an inverse association between LLIN use and distance to clinic with estimated net use dropping by more than 15% beyond a distance of 4km (not shown).

**Table 3:** Estimated risk ratios from univariate (left columns) and multivariate (right columns) log binomial regression modeling.

| Variable | RR | 95% CI | p-Value | aRR | 95% CI | p-Value |
| --- | --- | --- | --- | --- | --- | --- |
| **Elevation Quartile** |  |  |  |  |  |  |
| Quartile 1 |  | REF |  |  | REF |  |
| Quartile 2 | 1.06 | 1.03 – 1.10 | <0.001 | 1.07 | 1.03 – 1.11 | 0.02 |
| Quartile 3 | 1.01 | 0.97 – 1.04 | 0.73 | 0.97 | 0.93 – 1.02 | 0.23 |
| Quartile 4 | 0.86 | 0.83 – 0.89 | <0.001 | 0.87 | 0.81 – 0.94 | <.001 |
| **Distance to Clinic** |  |  |  |  |  |  |
| < 1 km |  | REF |  |  | REF |  |
| 1 - 2 km | 0.99 | 0.96 – 1.01 | 0.22 | 1.03 | 0.98 – 1.08 | 0.23 |
| ≥ 2 km | 0.86 | 0.83 – 0.89 | <0.001 | 0.73 | 0.64 – 0.82 | <.001 |

*Note: Univariate models separately regresses LLIN use on (1) elevation quartile and (2) distance to the nearest level II of level III health center. Multivariate model regresses LLIN use on elevation quartile and distance to the nearest level II of level III health center. Abbreviations: CI = confidence interval; km = kilometer; RR = risk ratio; aRR = adjusted risk ratio*

**Figure 2:**
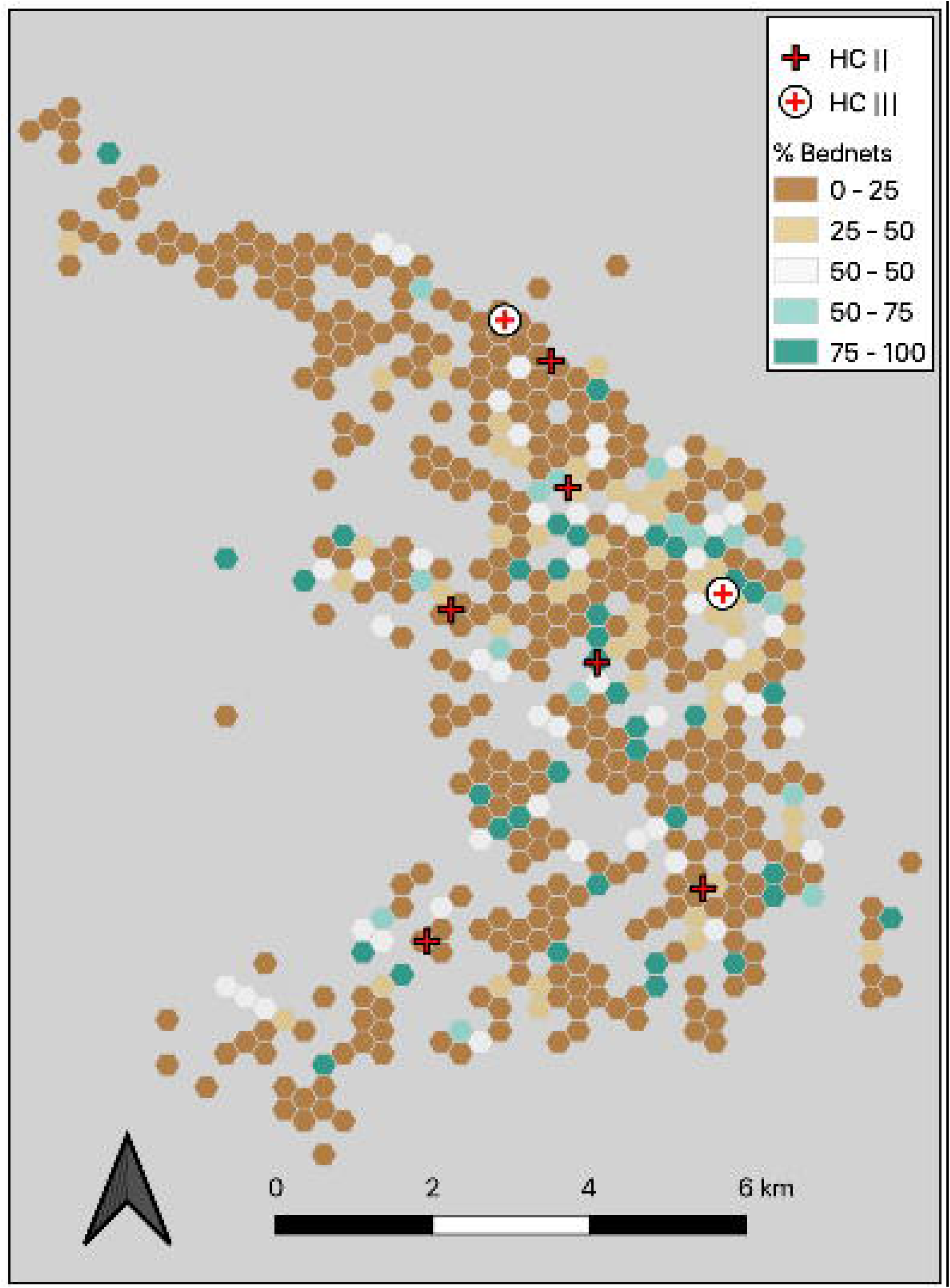
Map displaying the percentage of households that use a LLIN distributed through a health center compared to all households that use a bed net. Each hexagonal grid represents a minimal diameter of 200 meters.

Given that the majority of participants reported obtaining LLINs through a government campaign, we examined the association between geographic factors and net source. Overall, government mass distributions represented the primary source of nets across distance categories. However, households located farther from a health center were more likely to own nets sourced from mass distributions, while those located closer to health centers were more likely to own nets sourced through clinic visits (**Figure 3**). Those who received their LLINs from a mass distribution lived a median distance of 248 m (IQR 184 – 315 m, *p*<.001) farther from a health center than those who received a LLIN from a health facility.

**Figure 3:**
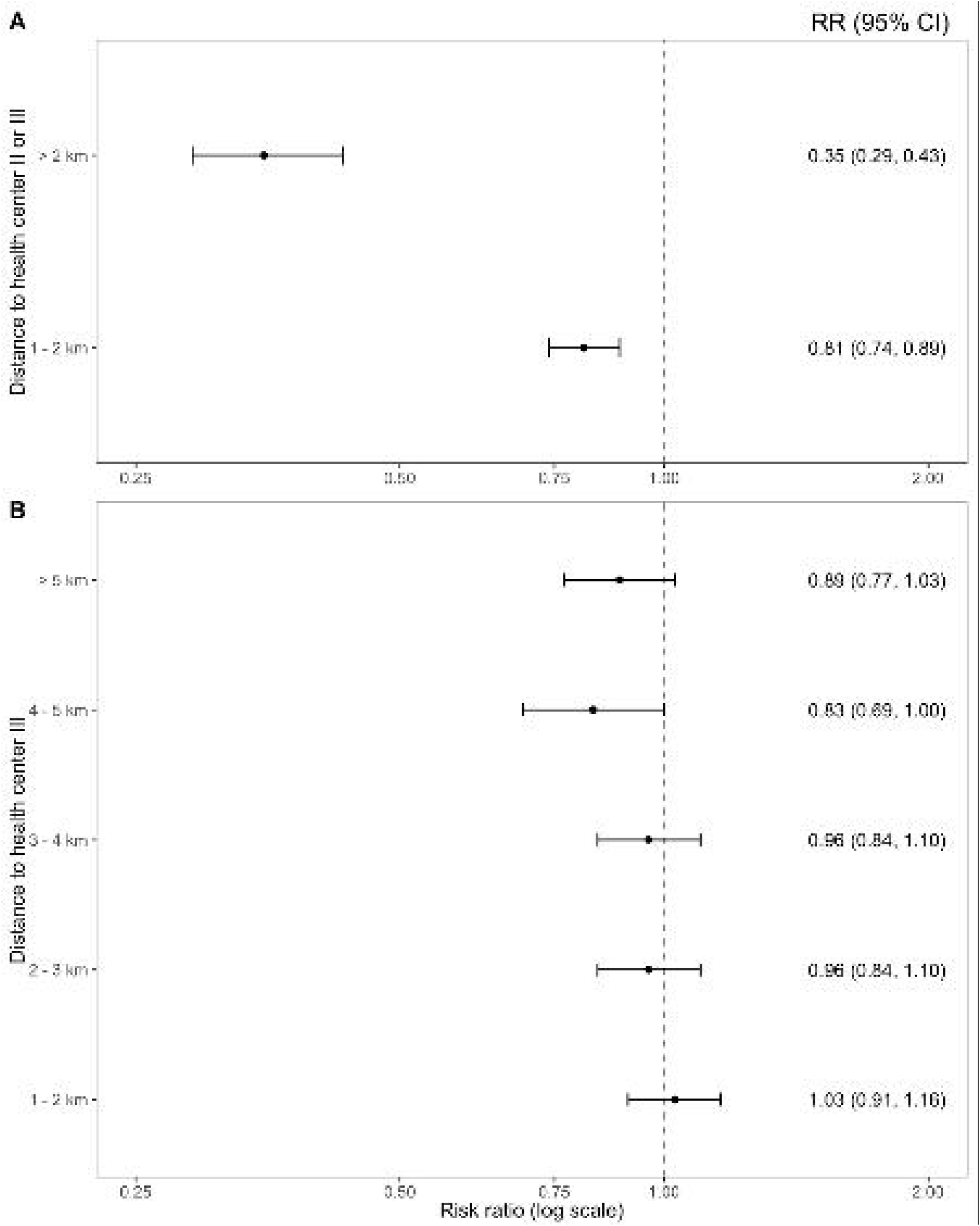
Estimated risk ratios of obtaining LLIN from health center (versus government distribution) by distance to health centers. Households living less than 1km from the health center are the reference group. *Note: Dropped observations where net was reported as “purchased” (n=4) or “other” (n=3)*.

## DISCUSSION

Despite Uganda’s substantial commitment to achieving universal LLIN coverage, our study - which was conducted near the end of a three-year government LLIN distribution cycle - suggests that approximately one-third of surveyed children did not sleep under a LLIN the previous night. The lowest rates of use were observed among households at elevations above 1,600 m and those farthest from health facilities. These results expand upon previous studies showing an inverse association between LLIN ownership or use and distance to a health facility, particularly in rural areas [23, 24]. Notably, households living ≥ 2 km from health facilities were much more likely to report receiving LLINs from mass distribution campaigns rather than from continuous distributions focused on high-risk patient populations at health facilities. While the observed differences in net sourcing are not unexpected, the findings do have important implications regarding implementation strategies to achieve and, perhaps more importantly, sustain universal LLIN coverage in rural Uganda.

While the WHO states that “mass campaigns are the only proven cost-effective way to rapidly achieve high and equitable coverage [7],” coverage gaps begin to appear almost immediately post-campaign due to net attrition well before the expiration manufacturer’s three-year lifespan [32-36]. Previous studies in Uganda have demonstrated the extent to which LLIN coverage and use declines in the interval period between distribution campaigns with only two-thirds of respondents reporting owning at least one LLIN three years after the last distribution campaign [12]. While we did not measure household coverage, our finding of 65% LLIN use by children (who are often more likely to sleep under nets) is consistent with these trends. Furthermore, declines in LLIN coverage and use have been associated with increased parasite prevalence, which highlights the need to develop novel strategies to replace lost and damaged nets between distribution campaigns [13].

Continuous distribution through existing health facilities is often cited as an effective supplemental strategy to overcome net attrition [7]. Yet our results suggest that this approach is insufficient to sustain coverage in rural areas, particularly as distance to health facilities increases with the greatest reduction in current use observed beyond a distance of 2 km. Households in these areas appear more dependent on mass distribution campaigns as the primary source of LLINs. In these more remote communities, school-based distributions may sustain higher and more equitable coverage [16, 17, 37]. Uganda also has an established network of community health workers, many of whom already perform evaluation and management of uncomplicated malaria, who could be leveraged to identify households without adequate LLINs [38-40]. This could take place through regular household surveys or as a component of febrile illness visits. Community-based malaria case management programs have been shown to reduce household costs associated with care seeking and similar benefits might be accrued if LLIN distribution was similarly decentralized [41].

The potential policy implications stemming from the finding of lower reported LLIN use at higher elevations, even when adjusting for distance to clinic, are more nuanced. We found that above 1,400 m the PfPR declines substantially with a number of the highest elevation villages having no positive RDT results; this result is consistent with the known association between malaria transmission and elevation [42, 43]. Furthermore, there does not appear to be any difference in the risk of malaria parasitemia between children sleeping under a net and those who did not (**Table 2**). Therefore, residents living at higher elevations may be making conscious decisions not to obtain or not to use LLINs given the lower risk of infection. Low perceived risk has been previously documented as a potential barrier to LLIN use [44]. While we did not assess travel histories or perform entomologic surveillance [45, 46], it is possible that some, if not most, of the infections identified at higher elevations may have been acquired during travel to lower-elevation market areas or social events (i.e., church, weddings). Given the lower prevalence of infection and minimal expected effect of LLINs on travel-related risk, our findings suggest that, at least from an economic standpoint, LLIN distribution at higher altitudes may be an inefficient use of resources. However, the additional effort and resources required to define discrete altitudinal thresholds at which LLIN distribution campaigns may no longer be effective may not be cost-effective, especially given that most of the Ugandan population resides well below these elevations. Furthermore, we acknowledge that despite potentially limited effectiveness for malaria control, LLIN distribution networks and distribution campaigns may serve other health and non-health goals, such as demonstrating the ability of local government to deliver essential services.

Our study, which was conducted in a setting of highly variable geography and malaria transmission intensity, has a number of strengths including the unique study area, the high-proportion of households sampled, and rigorous spatial mapping and analysis of individual households. There are also important limitations. First, we largely relied on self-reported outcomes such as bed net source and use. Participants may have perceived a social desirability pressure to state that the child had slept under the net. We are reassured that we observed differences in reported LLIN use across the elevation quartiles, as we would not expect a differential bias by elevation. Second, our use of RDTs may not have identified low-density (e.g., <50 parasites/µL), asymptomatic infections [27]. Given that RDTs are now widely employed for malaria indicator surveys, we believe this is a reasonable approach and is unlikely to have impacted our conclusions. Lastly, our site has large variations in elevation and malaria transmission intensity over a relatively small geographic area. While these characteristics make the site ideal for studies such as this, they may also limit the generalizability and utility of our findings to areas of more homogeneous terrain and/or transmission.

## CONCLUSIONS

In a setting of variable geography and malaria transmission, we found that continuous distribution efforts were insufficient to sustain high rates of LLIN use among children approximately three years after the last mass distribution campaign. Furthermore, geographic factors including elevation and distance to health facilities influenced reported rates of LLIN use. Households closer to health centers were more likely to have obtained a net from a health center, while households farther away were more likely to have a net from a government distribution and were less likely to use a net. Together, these findings suggest that more frequent mass distribution campaigns or combination implementation strategies may be required to achieve and maintain universal LLIN coverage.

## Data Availability

Deidentified individual data that supports the results will be shared beginning 9 to 36 months following publication provided the investigator who proposes to use the data has approval from an Institutional Review Board, Independent Ethics Committee, or Research Ethics Board, as applicable, and executes a data use/sharing agreement with UNC.

## DECLARATIONS

### Conflicts of Interests

All authors have completed the ICMJE uniform disclosure form and declare: no financial relationships with any organizations that might have an interest in the submitted work in the previous three years except that noted in the Funding section; no other relationships or activities that could appear to have influenced the submitted work.

### Previous Publication

The authors confirm that the results herein have not been previously presented or published and are not currently submitted or under review at another journal.

### Funding

The study was funded by the National Institutes of Allergy and Infectious Diseases (K23AI141764) awarded to RMB. VG acknowledges support from the Eunice Kennedy Shriver National Institute of Child Health and Human Development under award number P2C HD050924. The sponsor did not have any role in the design or conduct of the study or preparation of the manuscript.

### Author Contributions

Study conception and design: RMB, CC, BES, EMM. Funding: RMB, EM. Study implementation: RM, EB, MN, EMM, RMB. Analysis: CC, VG, BES, RMB. First draft of manuscript: CC, VG, BES, RMB. Revisions: All.

## Acknowledgements

We wish to thank the residents of the Bugoye sub-county, particularly the Village Health Teams, for their ongoing participation and support.

## ABBREVIATIONS

BHC: Bugoye Level III Health Center
C°: Degrees Celsius
CHW: Community Health Worker
CI: Confidence interval
DHS: Demographic and Health Survey
FPC: Finite population correction
HRPII: Histidine-rich protein II
IQR: Interquartile range
LLIN: Long-lasting insecticide-treated net
Km: Kilometer
M: Meter
PfPR: *Plasmodium falciparum* positivity rate
RDT: Rapid diagnostic test
RR: Risk ratio
SSA: sub-Saharan Africa

## Notes

### Competing Interest Statement

The authors have declared no competing interest.

